# Hidden changes to prespecified primary outcomes of clinical trials completed between 2009 and 2017 in German University Medical Centres: A meta-research study

**DOI:** 10.1101/2023.02.20.23286182

**Authors:** Martin R. Holst, Martin Haslberger, Samruddhi Yerunkar, Daniel Strech, Lars G. Hemkens, Benjamin G. Carlisle

## Abstract

**Objectives:** To assess how often clinical trials exhibit primary outcome discrepancies within registry records that would not be caught by comparing results publications to the latest registry entry, but would require analysing the registration history.

**Design:** Meta-research study.

**Setting:** All 1746 randomised clinical trials with published results, registered in ClinicalTrials.gov or Deutsches Register Klinischer Studien (DRKS), completed at German University Medical Centres between 2009 and 2017. We analysed registry entries for all trials and publications for a random sample of 292 trials.

**Participants:** Not applicable.

**Interventions:** Not applicable.

**Main outcome measures:** [1] Primary outcome discrepancies between registry entries at key study milestones and [2] the first results publication. [3] ‘Hidden’ discrepancies, i.e., only reported in the registry before the last entry, meaning they would only be detected by assessing the full registry change history. We considered discrepancies major if primary outcomes were newly added, dropped, or changed to or from secondary outcomes. [4] Proportion of publications transparently reporting discrepancies. [5] Characteristics associated with ‘open’ and ‘hidden’ discrepancies.

**Results:** Of all 1746 trials, 23% (n=393) had primary outcome discrepancies between trial start and latest registry entry, with 8% (n=142) being major. Primary outcomes in publications were different from the latest registry entry in 41% of trials (120 of the 292 sampled trials; 95% CI [35%, 47%]), with major discrepancies in 18% (54 of 292; 95% CI [14%, 23%]). ‘Hidden’ discrepancies were observed in 14% of trials (41/292; 95% CI [10%, 19%]). Only 1% of discrepancies were reported in the publications (2/161, 95% CI [0%, 4%]). Trials were more likely to have a within-registry discrepancy if they had an earlier registration year (OR 0.74; 95% CI [0.69, 0.80]), were registered on ClinicalTrials.gov (OR 0.41; 95% CI [0.23, 0.70]), or had been industry-sponsored (OR 0.29; 95% CI [0.21, 0.41]).

**Conclusions:** Changes to primary trial outcomes are common, often have major relevance, are rarely transparently reported and typically not detectable with an inspection of the latest registry entry. Authors need to be more transparent and registry entries of published trials need to require more in-depth analysis to reveal potentially misleading reporting practices.

**Protocol registration:** Open Science Framework (https://osf.io/t3qva; amendment in https://osf.io/qtd2b).

## Introduction

Clinical trial registries are critical tools to enhance the transparency and trustworthiness of clinical trial evidence by openly reporting key study parameters in a rapidly accessible manner (1). They make it possible to detect selective reporting biases that pose a major threat to trustworthiness. One of the most critical possible deviations from an original protocol and analysis plan is a change to prespecified outcomes. Consequently, outcomes and their pre-specification are among the most critical reporting items of clinical trials, with the CONSORT statement asking to describe ‘completely defined pre-specified primary and secondary outcome measures, including how and when they were assessed’ as well as ‘any changes to trial outcomes after the trial commenced, with reasons’ (2). If readers do not know if the outcomes were changed, they cannot assess the risk of bias due to ‘cherry picking’ of results or implications of multiple testing (3).

Many clinical trial registries, such as ClinicalTrials.gov, allow entries to be updated after initial registration. While updating a registry entry is generally useful, as it is not unusual for parts of a trial protocol to change and desirable for an entry to reflect the most current information, it constitutes another possible source of reporting bias if entries are not properly maintained and larger changes go unreported. While a number of ethical and reporting guidance documents recommend prospective trial registration (2,4,5) and some recommend the transparent reporting of protocol changes (2,4), this is only checked in a minority of cases by peer-reviewers (6). But even if considered, looking only at the latest registry entry may lead to a wrong assessment, as changes to the entry over the course of the study would remain ‘hidden behind the latest entry’. One would have to take a closer look at the history of changes, which some trial registries maintain, but which is often not easily accessible.

Several analyses have already assessed the frequency of outcome discrepancies between registries and publications. They widely agree that this problem is very common, with a median estimated prevalence of 31% of clinical trials affected by primary outcome changes (interquartile range across analyses: 17-45 %) (7). However, most studies published to date did not report which registry entry version they used (8–16), assessed discrepancies between the publication and the latest available registry entry (17–19), or the latest registry entry before trial completion (20); some also used the first available entry or first entry during the active phase (21,22). Some studies determined the timepoint of primary outcome registration and the timepoints of changes using registry histories (23–28). Only two studies assessed changes over the course of the registry history, using small samples and a simple terminology (29,30). No study quantified the number of changes missed by considering only the latest registry entries.

In this study, we aimed to assess [1] how often changes to primary outcomes of clinical trials are reported in clinical trial registries across all available registry entry versions (‘within-registry discrepancies’), [2] how often there are discrepancies between the latest registry entry and the results publication (‘registry-publication discrepancies’), [3] how many changes are ‘hidden’ behind the latest registry entry, i.e., are ‘within-registry’, but do not show up as ‘registry-publication discrepancies’ and are therefore easily missed in review, [4] how often both types of outcome changes are transparently reported in the results publications, and [5] which trial characteristics are associated with these reporting deficits.

## Methods

### Data sources and sample

We based our analyses on two published datasets (from the IntoValue projects, 31,32) containing all interventional studies completed at German University Medical Centres between 2009 and 2017. The trials had been registered in either ClinicalTrials.gov or the Deutsches Register Klinischer Studien (DRKS), which is the WHO primary trial registry for Germany. Both registries have an accessible history of changes. Our datasets also include links to corresponding results publications, which had been manually identified. We retrieved a combined dataset for both projects from a GitHub repository (https://github.com/maia-sh/intovalue-data), accessed on 24 January 2022).

### Eligibility criteria

We included any study that: 1) has a registry entry in either the ClinicalTrials.gov or the DRKS database, 2) was completed between 2009 and 2017 according to the trial status described as ‘Completed’, ‘Unknown status’, ‘Terminated’, or ‘Suspended’ (ClinicalTrials.gov), or ‘Recruiting complete, follow-up complete’, ‘Recruiting stopped after recruiting started’, or ‘Recruiting suspended on temporary hold’ (DRKS), 3) reported in the registry that a German University Medical Centre was involved (i.e., mentioned as responsible party, lead/primary sponsor, principal investigator, study chair, study director, facility, collaborator, or recruitment location; for definitions see 31,32), 4) has published results, i.e., a full-text publication of the trial results was found by the search methods described in the IntoValue protocols (31,32), 5) is registered as a randomised trial.

### Data extraction and processing

We used a stepwise approach of automatic data extraction and processing followed by reviewer assessment of identified outcome discrepancies, first comparing outcomes within the registry and second comparing registry entries with publications.

#### Automatic data extraction and processing

We obtained trial identifiers (NCT or DRKS number) and basic information about the trials and accompanying publications from the IntoValue dataset (31,32). We then downloaded all historical registry entries of these trials, using the cthist R package (33), on 24 January 2022 (DRKS) and 12 April 2022 (ClinicalTrials.gov). We extracted data from the fields listed in the codebook (https://osf.io/syux2). For our classification of within-registry discrepancies, we extracted primary and secondary outcomes at four key registration milestones: [1] the first entry after study start (inclusion of the first patient), [2] the latest entry before the end of active status (determined e.g. by ‘completed’ status in the registry), [3] the latest entry before publication of the results in a journal, and [4] latest available entry.

We classified trials into medical fields based on their journal’s category from the SCImago Journal & Country Rank database (34), which is based on the Scopus medical field classification. The journals’ categories were divided into 17 higher-order categories based on consensus (MRH, MH, LGH).

#### Assessment of within-registry outcome discrepancies

Two reviewers (MRH, MH) conducted the assessment of within-registry discrepancies in duplicate and resolved conflicts through discussion. Reviewers assessed changes to the primary outcomes between the four key trial milestones within the registry (Figure 1), using a classification system based on prior assessments of outcome switching (23,35) and descriptions of the specification of primary outcome measures (36) (Table 1).

**Table 1.** Categorisation of outcome discrepancies. Multiple categories could be present between two timepoints.

| **Major discrepancies** |  |
| --- | --- |
| New primary outcome | A new primary outcome was introduced in a registry entry. This also applied when (seemingly) composite outcomes or different timepoints were split into separate primary outcomes. |
| Primary from secondary | The new primary outcome was listed as a secondary outcome in the earlier registry entry. |
| Primary outcome omitted | A previously registered primary outcome was completely omitted in the later registry entry. |
| Primary to secondary | A previously registered primary outcome was reported as a secondary outcome in the later registry entry or publication. This also applied when (seemingly) composite outcomes were split into separate outcomes and some were later listed as secondary outcomes. |
| **Minor discrepancies** |  |
| Type of measurement changed, metric or method of aggregation changed, timing of assessment changed | Significant parts changed, e.g., timing from {24h} to {48h}. |
| Type of measurement specified, metric or method of aggregation specified, timing of assessment specified | Specified for the first time or significant detail added, e.g., from {seizure rate} to {seizure rate as recorded by family members}. |
| Type of measurement omitted, metric or method of aggregation omitted, timing of assessment omitted | Significant details of the registered primary outcomes were completely omitted or described with less specificity, e.g., from {seizure rate as recorded by family members} to {seizure rate}. |
| **No discrepancies** |  |
| No relevant change | We did not consider, for example:<br>• Correction of typos,<br>• description of statistical analyses (e.g., intention-to-treat vs per-protocol population, statistical tests used),<br>• description of handling of missing data,<br>• addition of redundant descriptions of known scales (e.g., RECIST criteria, Visual Analogue Scale), or<br>• redundant details pertaining to measurement (e.g., change from baseline). |

**Figure 1.**
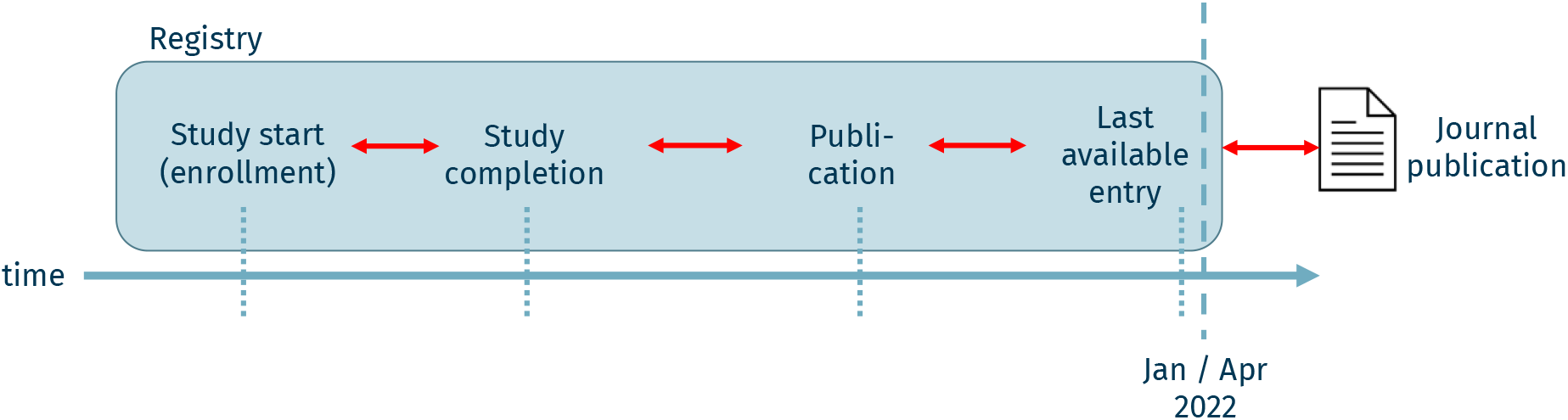
Key trial timepoints analysed for outcome discrepancies in this study. Any changes of primary outcomes between study start and last available entry (i.e., within-registry discrepancies; registry data were downloaded in January 2022 for DRKS and in April 2022 for ClinicalTrials.gov) were considered and compared to the journal publication (i.e., registry-publication discrepancies).

#### Assessment of registry-publication outcome discrepancies

For feasibility, we drew a random sample of 300 publications, from which three reviewers (MRH, MH, SY) extracted primary outcomes. Some of these publications, however, turned out to not be the primary results publications of their respective trials. If the reviewers could not find the correct publication, they excluded the publication, reducing the final sample size to 292. We considered outcomes primary if they were explicitly named as such using a list of keywords (‘primary’ / ‘main’ / ‘outcome’ / ‘endpoint’ / ‘end point’). Otherwise, we used the outcome used for sample size calculation. In all other cases, we used the first reported outcome in the abstract and results section (with priority given to the abstract). For each entry, reviewers compared the extracted primary outcome(s) with the latest available registry entry using the developed rating system (Table 1). Reviewers also recorded whether there were any mentions of outcome changes in the assessed publications. Conflicts were resolved through discussion between raters.

#### Inter-rater agreement

The overall inter-rater agreement over all timepoints and items (measured by Cohen’s kappa) during screening was κ = 0.81 for the within-registry ratings and κ = 0.46 during the registry-publication ratings. Disagreement resulted partly from cases without clear reporting of primary outcomes in publications (if only explicitly mentioned outcomes were used for the registry-publication ratings, inter-rater agreement was κ = 0.51). In a post-hoc sensitivity analysis we restricted our analysis to primary outcomes explicitly named as such in the publications or used for sample size calculation.

### Statistical analyses and reporting

We report descriptive statistics for the rates of different types of outcome discrepancies within the registry entries (between key trial milestones), and between the latest registry entries and results publications. For proportions within the random sample of 292 trials with manual assessment of publications, we report 95% confidence intervals. For the analysis of determinants of within-registry outcome discrepancies or registry-publication outcome discrepancies, we used logistic regressions, with either any within-registry outcome discrepancy, or any registry-publication outcome discrepancy, as response variables, and a set of 9 candidate predictors (study phase, sponsor, publication year, registration year, medical field, registry, multicenter trial, enrollment, intervention). These variables were prespecified before the start of regression analyses (details in Supplementary Table 1).

#### Software

We used the R package cthist (33) to download clinical trial registration histories, Numbat Systematic Review Manager (37) for data extraction, and R (38) for data processing and statistical analyses.

#### Reporting

We used the STROBE guideline (39) to structure our manuscript.

## Results

We included 1746 clinical trials completed at German University Medical Centres between 2009 and 2017 (Table 2; Figure 2). Median registered sample size of these trials was 100, their median registration year 2011 and median publication year 2015. The largest fields represented by these trials were general medicine (29%) and internal medicine (17%). Industry was the lead sponsor for 26% of trials.

**Table 2.**
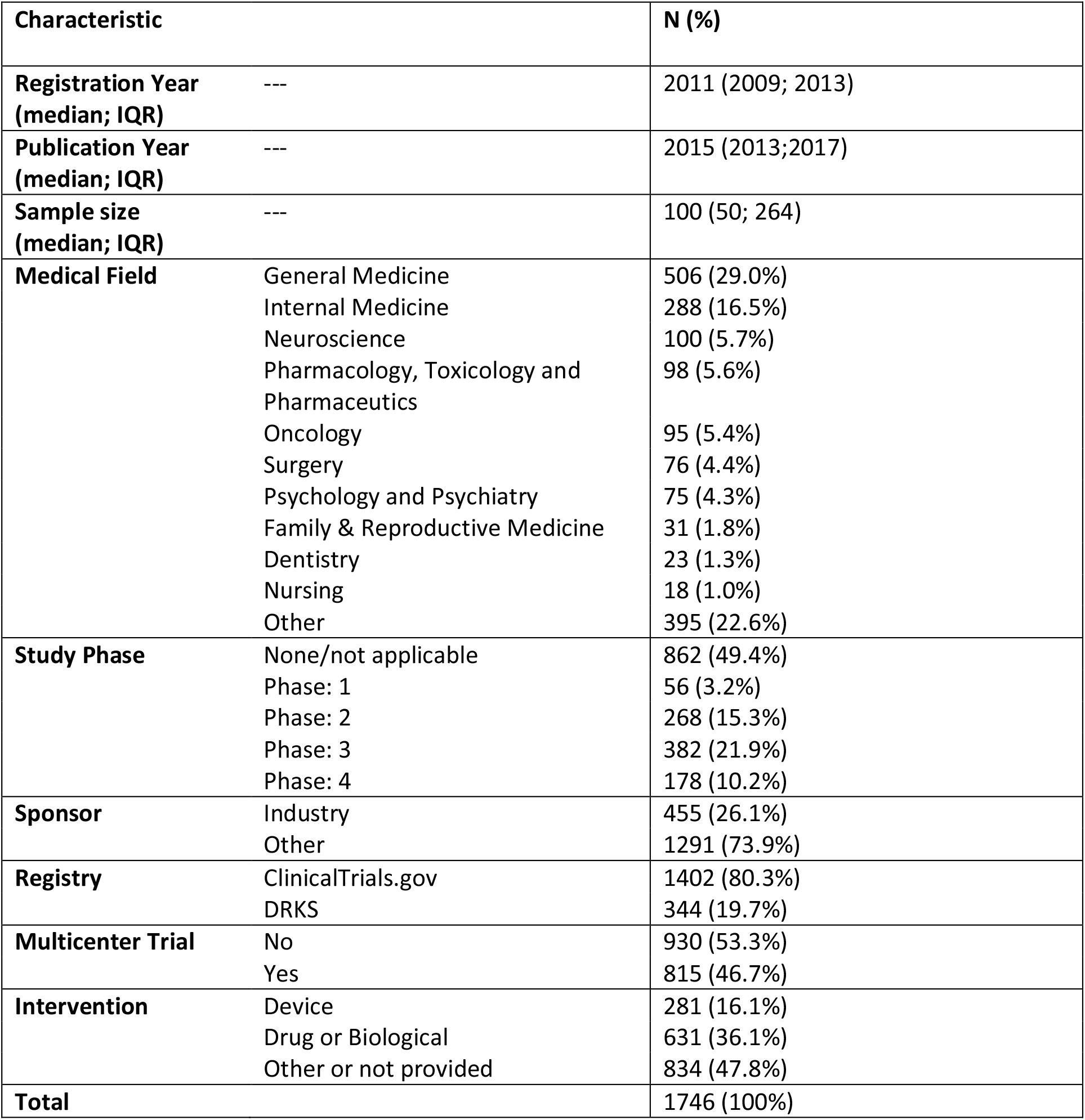
Characteristics of included trials.

| Characteristic |  | N (%) |
| --- | --- | --- |
| **Registration Year (median; IQR)** | --- | 2011 (2009; 2013) |
| **Publication Year (median; IQR)** | --- | 2015 (2013;2017) |
| **Sample size (median; IQR)** | --- | 100 (50; 264) |
| **Medical Field** | General Medicine<br>Internal Medicine<br>Neuroscience<br>Pharmacology, Toxicology and<br>Pharmaceutics<br>Oncology<br>Surgery<br>Psychology and Psychiatry<br>Family & Reproductive Medicine<br>Dentistry<br>Nursing<br>Other | 506 (29.0%)<br>288 (16.5%)<br>100 (5.7%)<br>98 (5.6%)<br>95 (5.4%)<br>76 (4.4%)<br>75 (4.3%)<br>31 (1.8%)<br>23 (1.3%)<br>18 (1.0%)<br>395 (22.6%) |
| **Study Phase** | None/not applicable<br>Phase: 1<br>Phase: 2<br>Phase: 3<br>Phase: 4 | 862 (49.4%)<br>56 (3.2%)<br>268 (15.3%)<br>382 (21.9%)<br>178 (10.2%) |
| **Sponsor** | Industry<br>Other | 455 (26.1%)<br>1291 (73.9%) |
| **Registry** | ClinicalTrials.gov<br>DRKS | 1402 (80.3%)<br>344 (19.7%) |
| **Multicenter Trial** | No<br>Yes | 930 (53.3%)<br>815 (46.7%) |
| **Intervention** | Device<br>Drug or Biological<br>Other or not provided | 281 (16.1%)<br>631 (36.1%)<br>834 (47.8%) |
| **Total** |  | 1746 (100%) |

**Figure 2.**
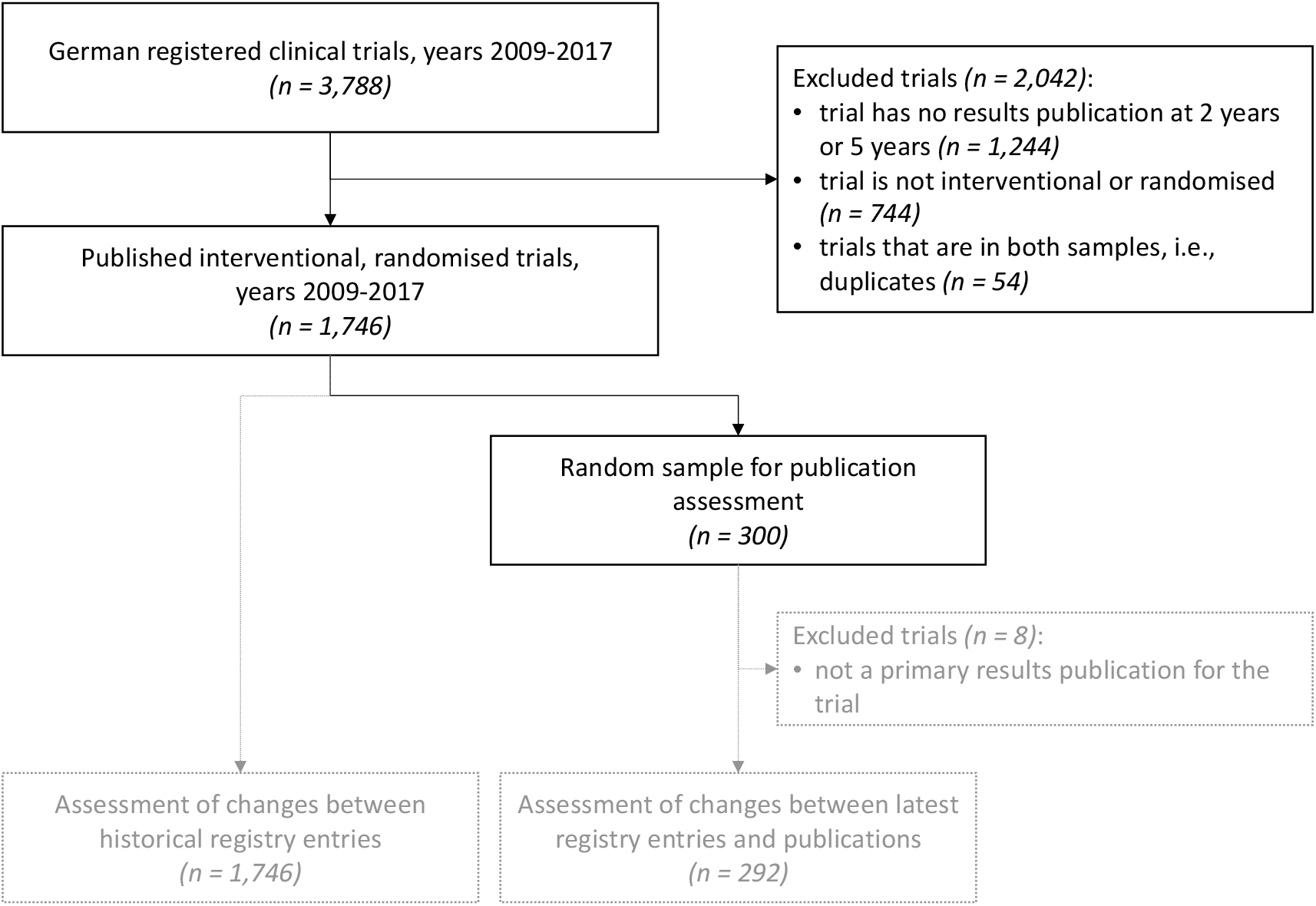
Study flowchart.

### Prevalence of outcome discrepancies

#### Primary outcome discrepancies within trial registries

Of 1746 trials, 393 (23%) had an outcome discrepancy reported within the registry (Table 3; Figure 3), 142 trials (8%) had major discrepancies, i.e., a primary outcome was newly added, dropped, or changed to or from a secondary outcome. Of these major discrepancies, 66 (4%) happened during the active phase of the trial, i.e., after first patient inclusion and before the trial status was set to ‘completed’, another 49 (3%) between completion and publication, and 36 (2%) after publication. Minor discrepancies were seen in 318 (18%) trials.

**Table 3.** Primary outcome discrepancies in 1746 trials published between 2009 to 2017. The table shows the changes of primary outcomes reported in the registries at different trial timepoints compared to previous registry entries and the discrepancies detected in results publications compared to the latest registry entry.

|  | Within-registry discrepancies per trial timepoint (n=1746) |  |  |  | Registry-publication<br>discrepancies<br>(n=292; sample) |
| --- | --- | --- | --- | --- | --- |
|  | Any | recruitment | post-<br>completion | post-<br>publication |  |
| Discrepancies<br>**any** | **393 (22.51%)** | **167 (9.56%)** | **159 (9.11%)** | **131 (7.50%)** | **120 (41.10%)**<br>**[35.40%, 46.98%]** |
| major | 142 (8.13%) | 66 (3.78%) | 49 (2.81%) | 36 (2.06%) | 54 (18.49%)<br>[14.21%, 23.43%] |
| minor | 318 (18.21%) | 117 (6.70%) | 130 (7.45%) | 110 (6.30%) | 75 (25.68%)<br>[20.77%, 31.10%] |
| - changes | 149 (8.53%) | 49 (2.81%) | 61 (3.49%) | 51 (2.92%) | 45 (15.41%)<br>[11.47%, 20.07%] |
| - addition/omission | 233 (13.34%) | 78 (4.47%) | 91 (5.21%) | 80 (4.58%) | 32 (10.96%)<br>[7.62%, 15.12%] |
| **milestone does not**<br>**exist** | **5 (0.29%)** | **332 (19.01%)** | **266 (15.23%)** | **945 (54.12%)** | **0 (0.00%)**<br>**[0.00%, 0.00%]** |
| **none** | **1348 (77.21%)** | **1247 (71.42%)** | **1321 (75.66%)** | **670 (38.37%)** | **172 (58.90%)**<br>**[53.02%, 64.60%]** |

**Figure 3.**
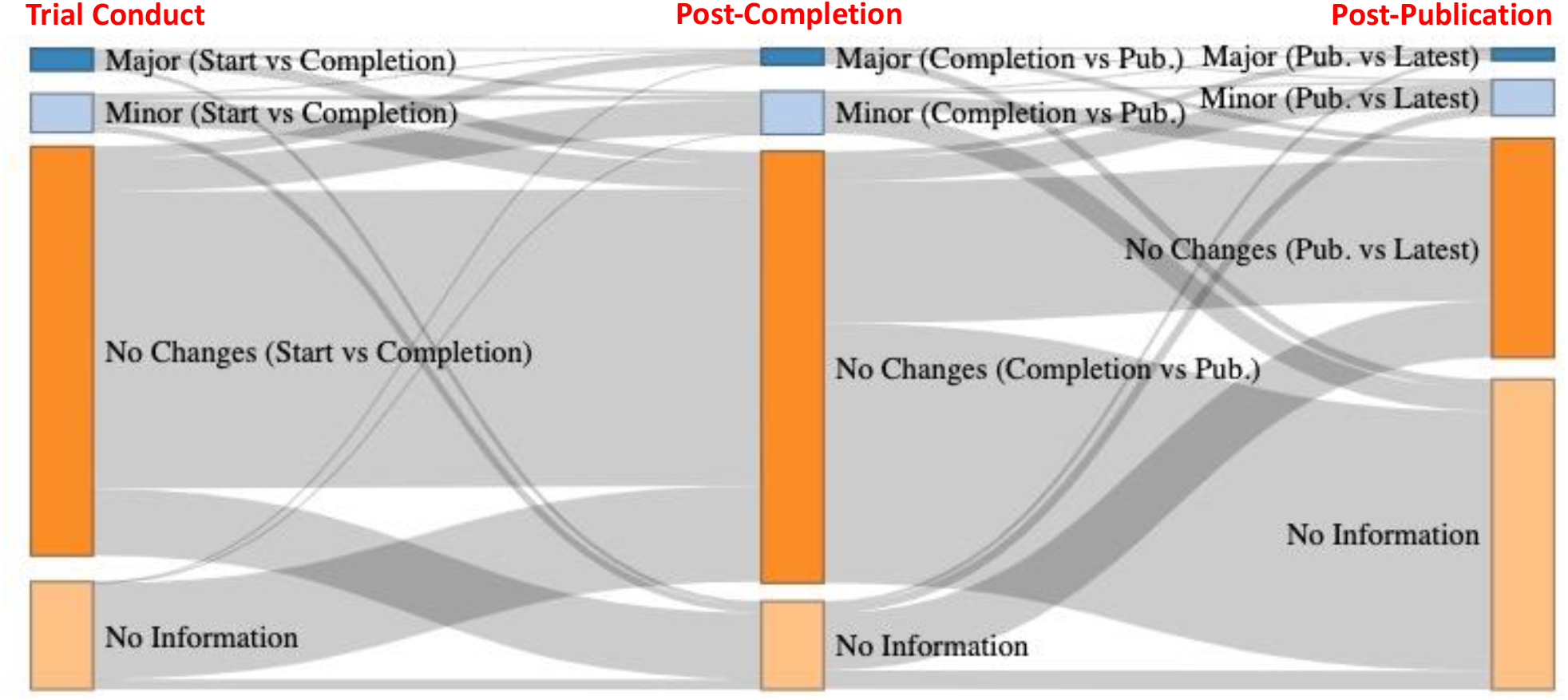
Percentage of trials showing primary outcome discrepancies within the registry history per trial phase. ‘No information’ means that a trial did not have a registry entry at that time or did not have any updates to the registry entry at that time.

#### Primary outcome discrepancies between registry entries and publications

Out of 292 randomly selected trials among the 1746 trials, 120 (41%; 95% CI [35%, 47%]) had a discrepancy between the latest registry entry and the publication, of which 54 (18%; 95% CI [14%, 23%]) were major and 75 (26%; 95% CI [21%, 31%]) were minor discrepancies.

### Hidden changes of primary outcomes

Among the 292 trials, 161 (55%: 95% CI [49%, 61%]) had an outcome discrepancy at any stage (Figure 4; Figure S1 and S2). There were discrepancies only between the latest registry entry and the publication in 95 trials (33%; 95% CI [27%, 38%]), with 42 being major (14%) and 61 being minor (21%); discrepancies only within the registry histories in 41 trials (14%; 95% CI [10%, 19%]) with 13 being major (4%) and 35 being minor (12%); and 25 trials (9%; 95% CI [6%, 12%]), 12 major (4%) and 14 minor (5%), had discrepancies both between the latest registry entry and the publication and also within the registry.

**Figure 4.**
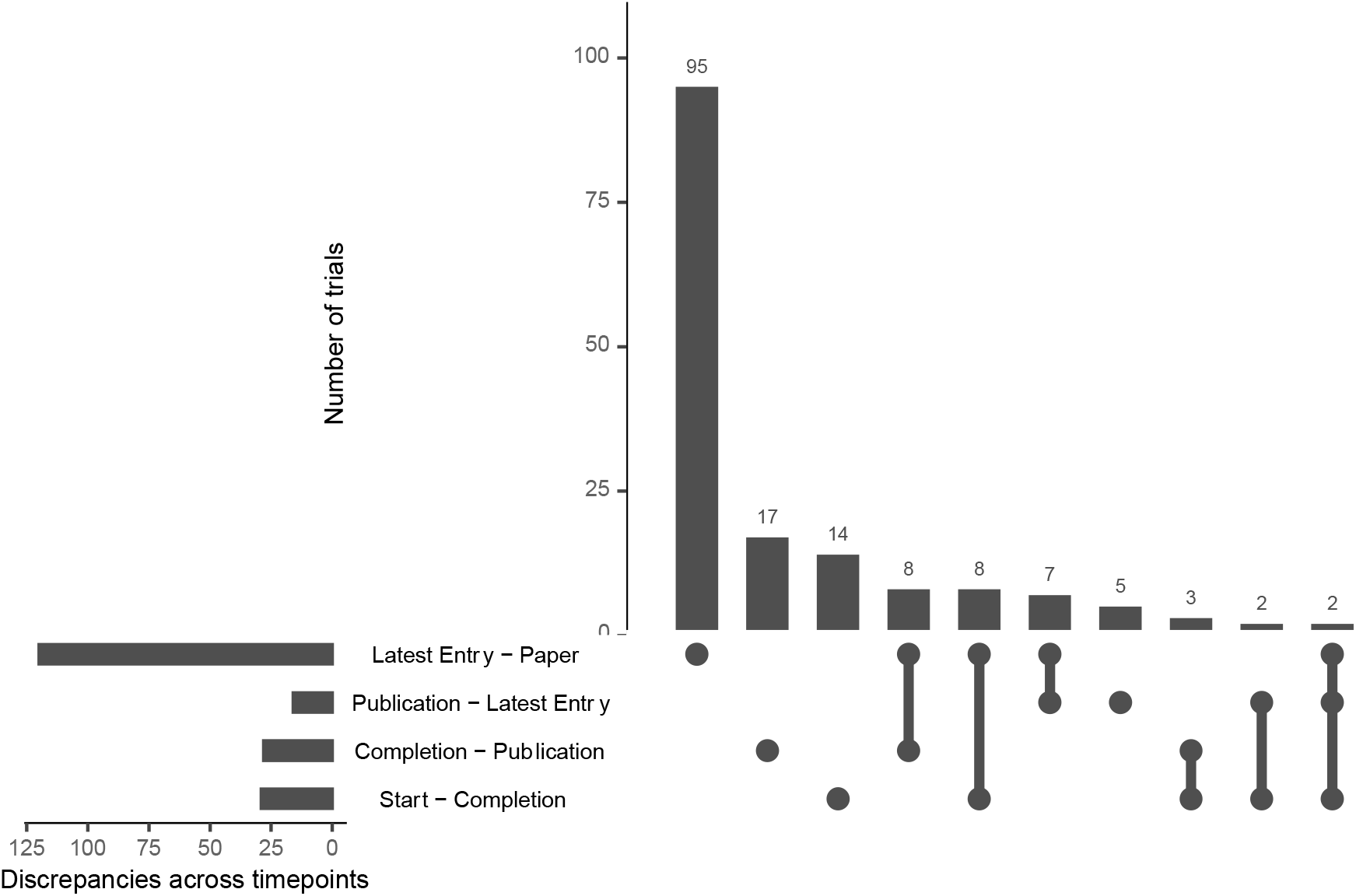
UpSet plot showing the prevalence of any change at the different time point combinations in a random sample of 292 trials. The first bar from the left shows the number of trials that only show a discrepancy between the latest entry and the publication (95 trials, 32.5%). The number of trials showing outcome discrepancies within the registry at different timepoints after trial start (all other bars), adds up to 66 trials (22.6%). 41 trials (14.0%) exhibit changes in the registry, but the latest entry and outcome description in the publication correspond, i.e., outcome discrepancies would not be detected by comparing the publication to the latest available registry entry. 131 trials did not have any change in the registry or between registry and publication (44.9%).

### Reporting of changes in the publications

Only two of the 161 trials with any change to the outcomes reported this change in the publication (1%, 95% CI [0%, 4%]). One of the trials, which exhibited a major change both within the registry (between completion and publication) and between the latest registry entry and publication, stated that some of the primary outcomes were going to be reported in a separate manuscript (40). The other publication, which had a major change between the latest entry and the publication, stated that the changed outcome was the ‘more sensitive and direct measurement’ (41).

### Trial characteristics associated with outcome changes (within-registry and registry-publication)

Within-registry outcome changes were more likely in industry-sponsored trials (47% Industry vs 14% Non-Industry, OR 0.29, 95% CI [0.21, 0.41], p < 0.001; Supplementary Table 1), trials registered on ClinicalTrials.gov (27% ClinicalTrials.gov vs 5% DRKS, OR 0.41, 95%CI [0.23, 0.70], p = 0.002), and trials that were registered earlier (per registration year; OR 0.74, 95% CI [0.69, 0.80], p < 0.001). No statistically significant association with any other trial characteristic was found.

For registry-publication outcome discrepancies, we identified no statistically significant association with any candidate predictor (Supplementary Table 2).

We detected no statistically significant association between within-registry outcome discrepancies and registry-publication outcome discrepancies (Supplementary Table 3).

Results were similar in the sensitivity analysis (Supplementary Table 4).

## Discussion

Our assessment of all 1746 published clinical trials conducted at German university medical centres that were completed between 2009 and 2017 shows that changes to primary outcomes are very common. About one in four trials (23%) has discrepancies within the registry after study start; a major discrepancy occurs within one in twelve trials (8%). Of the 292 assessed publications, 41% have discrepancies between latest registry entry and the outcomes presented to readers, and 14% of the trials had changes ‘hidden’ within the registry, with no visible discrepancy between the latest entry and the publication. These ‘hidden’ discrepancies were major in 4% of trials. An inspection of only the latest registry entry alongside the publication, without assessment of the entire registration history, would miss a primary outcome change, which is often major, in one in seven trials. We used a comparatively large sample of German clinical trials registered between 2009 and 2017, with a validated set of corresponding results publications for registry entries, determined by an extensive manual screening process (31,32). Our methodology allowed us to detect changes to registered primary outcomes that might have gone unnoticed by other studies, by retrieving all historical versions for trials, and detecting changes using a streamlined workflow. This is different from previous studies, which instead used manual searches (29,30). Many trials in our sample had changes to the registry after study completion (9%) or after publication of the first results paper (8%). The changes were almost never reported in the paper. We revealed potential indicators for these practices, with trials that were registered earlier, trials registered on ClinicalTrials.gov, and industry-sponsored trials having a higher likelihood of within-registry outcome discrepancies.

Assessments of outcome changes that rely on a single version of registry entries might underestimate the true prevalence of discrepancies. The ICMJE policy states that any changes to the registration should be explained by authors in the publication (4). Both the observed lack of reporting on outcome discrepancies in results publications, as well as survey results among manuscript reviewers (6), indicate that reviewers check the registry entry of a clinical trial only in the minority of cases. While registries make change histories available and offer tools to compare different registration versions, and journal editors and reviewers in many cases use these tools to check the registration histories, the low number of reporting of discrepancies in primary outcomes suggests that this is not properly enforced. To counteract, journals could implement editorial policies that do require editors, peer reviewers, or other personnel to assess the historical versions of submitted clinical trials. Clinical trial registries, on the other hand, could help by implementing solutions to identify and mark trials with major discrepancies in their outcomes.

Still, some trials change their outcomes at two or even three different timepoints over the course of the trial. Interestingly, we found not only cases in which primary outcomes gained more detail, but also cases in which primary outcomes were described with less detail than before. Overall, we found 55% of trials to have some form of outcome change over the course of the study. This is somewhat higher compared to other studies, which exhibit a large range for the frequency, with a median of 31% (Interquartile Range: 17-45 % (7)). This might be due to different methodology, samples, timepoints, or definition of outcome discrepancies. A common reference used in other assessments of outcome discrepancies is the study by Chan et al. (35) that used a more narrow definition of outcome changes (roughly corresponding to our ‘major’ category). We used a relatively liberal definition of primary outcome changes, defining added or omitted specificity as minor discrepancies. This study has several limitations. First, we rely on the reporting quality of published studies. In 48 out of 292 publications, the primary outcome was not explicitly mentioned in the publication, so we had to take the outcome used for sample size calculation or the first reported outcome instead. A sensitivity analysis excluding these 48 studies supported our main analysis. Second, the categorisation of outcome changes is somewhat subjective, and while the agreement was very high for the within-registry changes (κ = 0.81), it was moderate (κ = 0.46) for the registration-publication ratings due to the added complexity of identifying the correct outcome as ‘primary’. However, both conflicting primary outcome definitions and conflicting ratings were resolved in discussions between the reviewers, and excluding unclear situations in the abovementioned sensitivity analysis did not change the main findings. Third, although we used high quality underlying data reflecting German clinical research output, we only assess a single geographic area, partly reflecting EU/German registration policies. Finally, we only looked at changes between registry entries but did not check how many outcome measures that show no change remain underspecified (e.g., no information on the type of measurement, timing, or method of aggregation).

Overall, our analysis did not only reveal these research findings but also demonstrated the feasibility of an efficient workflow (33) that can be used for future projects and overcome previously described challenges to incorporate historical registration data (42).

## Conclusion

Primary outcome discrepancies are very common in clinical research. Such discrepancies are often hidden in the trial registries and almost never reported in publications. The problem seems to be ubiquitous, but may be more common in older trials, trials registered in ClinicalTrials.gov, and industry-sponsored trials. Even though trial registries can be an important tool to detect outcome discrepancies, review procedures employed by journals seem to rarely make use of them. Our approach provides a feasible approach to further investigate this issue. Overall, it is not sufficient to only assess the publication and latest registry entry – a careful assessment of the full research conduct from conception to publication is needed to ensure trustworthy evidence for clinical decisions.

## Data Availability

Analysis code and original datasets are available on Github (https://github.com/Martin-R-H/HiddenOutcomeChanges). The final dataset has been posted to our OSF page (https://osf.io/e2uct/).

