## Supplementary Materials for "Hidden changes to prespecified primary outcomes of clinical trials completed between 2009 and 2017 in German University Medical Centres: A meta-research study"

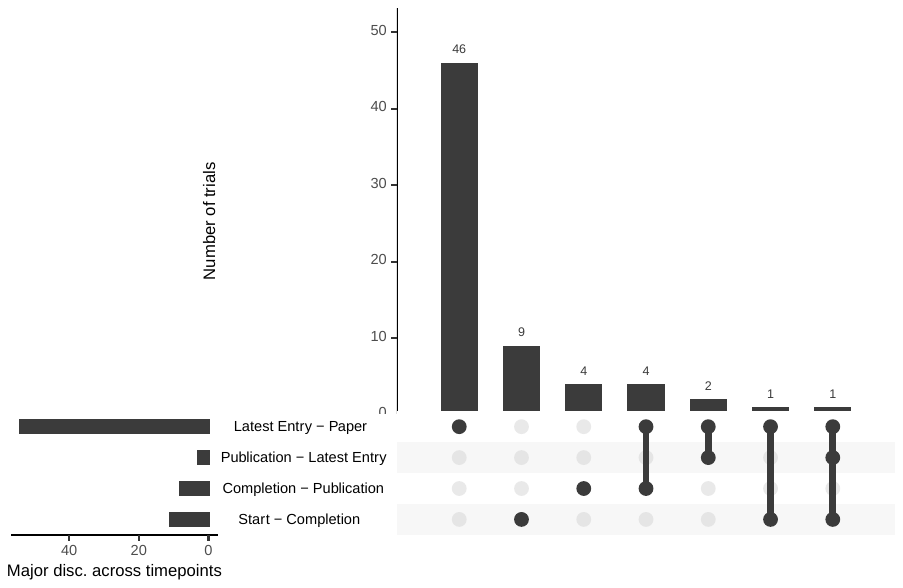

*Figure S1.* UpSet plot showing the prevalence of major discrepancies at the different time point combinations in a random sample of 292 trials. The first bar from the left shows the number of trials that only show a discrepancy between the latest entry and the publication (46 trials, 15.8%). The number of trials showing major outcome discrepancies within the registry at different timepoints after trial start (all other bars), adds up to 21 trials (7.2%). 13 trials (4.5%) exhibit major discrepancies in the registry, but the latest entry and outcome description in the publication correspond, i.e., major outcome discrepancies would not be detected by comparing the publication to the latest available registry entry. 225 trials did not have major discrepancies in the registry or between registry and publication (77.1%).

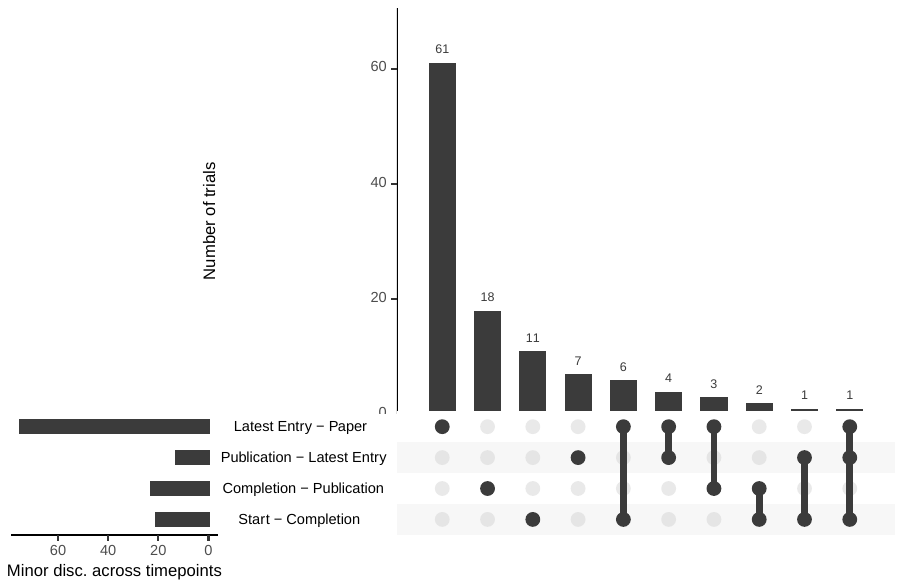

*Figure S2.* UpSet plot showing the prevalence of minor discrepancies at the different time point combinations in a random sample of 292 trials. The first bar from the left shows the number of trials that only show a discrepancy between the latest entry and the publication (61 trials, 20.9%). The number of trials showing minor outcome discrepancies within the registry at different timepoints after trial start (all other bars), adds up to 53 trials (18.2%). 39 trials (13.4%) exhibit minor discrepancies in the registry, but the latest entry and outcome description in the publication correspond, i.e., minor outcome discrepancies would not be detected by comparing the publication to the latest available registry entry. 178 trials did not have minor discrepancies in the registry or between registry and publication (61.0%).

*Supplementary Table 1.* Regression coefficients and p-values for our logistic regression model, with any within-registry outcome change as the response variable. None of the predictors had missing data.

| **Predictor** | **Predictor Level** | **Any outcome discrepancy** | **No outcome discrepancy** | **Odds Ratio (exponential of regression coefficient)** | **p-Value** |
| --- | --- | --- | --- | --- | --- |
| **Study Phase** | No phase (Intercept) | 113 (13.1) | 749 (86.9) |  |  |
|  | Phase: 1 | 6 (10.7) | 50 (89.3) | 0.68 | 0.44 |
|  | Phase: 2 | 75 (28.0) | 193 (72.0) | 0.71 | 0.16 |
|  | Phase: 3 | 155 (40.6) | 227 (59.4) | 1.045 | 0.84 |
|  | Phase: 4 | 44 (24.7) | 134 (75.3) | 0.81 | 0.38 |
| **Sponsor** | Industry | 212 (46.6) | 243 (53.4) |  |  |
|  | Other | 181 (14.0) | 1110 (86.0) | 0.29 | < 0.0001* (0.0000000000001723) |
| **Publication Year** | --- | 2014.0 (2.6) | 2015.3 (2.5) | 1.06 | 0.07 |
| **Registration Year** | --- | 2009.3 (2.9) | 2011.9 (2.8) | 0.74 | < 0.0001* (< 0.0000000000000002) |
| **Medical Field** | Basic (Intercept) | 1 (14.3) | 6 (85.7) |  |  |
|  | Dentistry | 3 (13.0) | 20 (87.0) | 0.87 | 0.91 |
|  | Epidemiology and Public Health | 3 (30.0) | 7 (70.0) | 1.91 | 0.63 |
|  | Family & Reproductive Medicine | 6 (19.4) | 25 (80.6) | 0.47 | 0.54 |
|  | General Medicine | 126 (24.9) | 380 (75.1) | 0.63 | 0.68 |
|  | Health Professions | 0 (0.0) | 9 (100.0) | < 0.01 | 0.98 |
|  | Immunology and Microbiology | 4 (26.7) | 11 (73.3) | 0.56 | 0.66 |
|  | Internal Medicine | 79 (27.4) | 209 (72.6) | 0.66 | 0.71 |
|  | Neuroscience | 18 (18.0) | 82 (82.0) | 0.52 | 0.57 |
|  | Nursing | 2 (11.1) | 16 (88.9) | 0.89 | 0.93 |
|  | Oncology | 41 (43.2) | 54 (56.8) | 1.37 | 0.78 |
|  | Other | 63 (19.3) | 264 (80.7) | 0.71 | 0.76 |
|  | Other Clinical Field | 10 (27.8) | 26 (72.2) | 0.49 | 0.55 |
|  | Other Medical Field | 2 (6.2) | 30 (93.8) | 0.57 | 0.67 |
|  | Pharmacology, Toxicology and Pharmaceutics | 15 (15.3) | 83 (84.7) | 0.71 | 0.77 |
|  | Psychology and Psychiatry | 6 (8.0) | 69 (92.0) | 0.42 | 0.47 |
|  | Surgery | 14 (18.4) | 62 (81.6) | 0.54 | 0.60 |
| **Registry** | ClinicalTrials.gov (Intercept) | 375 (26.7) | 1027 (73.3) |  |  |
|  | DRKS | 18 (5.2) | 326 (94.8) | 0.41 | < 0.01* (0.00169) |
| **Multicenter Trial** | No (Intercept) | 120 (12.9) | 810 (87.1) |  |  |
|  | Yes | 273 (33.5) | 542 (66.5) | 1.22 | 0.22 |
| **Enrollment** | --- | 387.6 (613.2) | 280.2 (1126.1) | 1.00 | 0.30 |
| **Intervention** | Device (Intercept) | 81 (28.8) | 200 (71.2) |  |  |
|  | Drug or Biological | 221 (35.0) | 410 (65.0) | 0.83 | 0.39 |
|  | Other | 91 (10.9) | 743 (89.1) | 0.78 | 0.25 |

*Supplementary Table 2.* Regression coefficients and p-values for our logistic regression model, with any registry-publication outcome change as the response variable (n = 292 trials). None of the predictors had missing data.

| **Predictor** | **Predictor Level** | **Any outcome change** | **No outcome change** | **Odds Ratio (exponential of regression coefficient)** | **p-Value** |
| --- | --- | --- | --- | --- | --- |
| **Study Phase** | No phase (Intercept) | 72 (46.8) | 82 (53.2) |  |  |
|  | Phase: 1 | 0 (0.0) | 6 (100.0) | < 0.01 | 0.99 |
|  | Phase: 2 | 16 (42.1) | 22 (57.9) | 1.16 | 0.78 |
|  | Phase: 3 | 20 (32.2) | 42 (67.7) | 0.58 | 0.23 |
|  | Phase: 4 | 12 (37.5) | 20 (62.5) | 0.76 | 0.56 |
| **Sponsor** | Industry | 27 (39.1) | 42 (60.9) |  |  |
|  | Other | 93 (41.7) | 130 (58.3) | 0.75 | 0.46 |
| **Publication Year** | --- | 2015.0 (2.9) | 2015.3 (2.5) | 0.96 | 0.56 |
| **Registration Year** | --- | 2011.3 (3.0) | 2011.5 (2.9) | 0.92 | 0.27 |
| **Medical Field** | Basic (Intercept) | 0 (0.0) | 1 (100.0) |  |  |
|  | Dentistry | 2 (66.7) | 1 (33.3) | 18712225.43 | 0.99 |
|  | Epidemiology and Public Health | 3 (100.0) | 0 (0.0) | 219310419443773.03 | 0.99 |
|  | Family & Reproductive Medicine | 3 (50.0) | 3 (50.0) | 20784418.13 | 0.99 |
|  | General Medicine | 32 (40.0) | 48 (60.0) | 9417882.88 | 0.99 |
|  | Immunology and Microbiology | 1 (100.0) | 0 (0.0) | 159072914643630.18 | 0.99 |
|  | Internal Medicine | 21 (38.9) | 33 (61.1) | 8108902.18 | 0.99 |
|  | Neuroscience | 7 (50.0) | 7 (50.0) | 11814401.98 | 0.99 |
|  | Nursing | 0 (0.0) | 1 (100.0) | 0.76 | 1.00 |
|  | Oncology | 5 (33.3) | 10 (66.7) | 8857066.81 | 0.99 |
|  | Other | 24 (40.0) | 36 (60.0) | 7898684.53 | 0.99 |
|  | Other Clinical Field | 2 (66.7) | 1 (33.3) | 25094737.46 | 0.99 |
|  | Other Medical Field | 1 (25.0) | 3 (75.0) | 5407042.91 | 0.99 |
|  | Pharmacology, Toxicology and Pharmaceutics | 4 (21.1) | 15 (78.9) | 3577545.31 | 1.00 |
|  | Psychology and Psychiatry | 7 (53.8) | 6 (46.2) | 15284871.01 | 0.99 |
|  | Surgery | 8 (53.3) | 7 (46.7) | 11098490.54 | 0.99 |
| **Registry** | ClinicalTrials.gov (Intercept) | 92 (38.3) | 148 (61.7) |  |  |
|  | DRKS | 28 (53.8) | 24 (46.2) | 1.81 | 0.12 |
| **Multicenter Trial** | No (Intercept) | 74 (47.7) | 81 (52.3) |  |  |
|  | Yes | 46 (33.6) | 91 (66.4) | 0.56 | 0.08 |
| **Enrollment** | --- | 202.4 (308.9) | 298.8 (580.1) | 1.00 | 0.24 |
| **Intervention** | Device (Intercept) | 21 (44.7) | 26 (55.3) |  |  |
|  | Drug or Biological | 32 (33.3) | 64 (66.7) | 0.62 | 0.29 |
|  | Other | 67 (45.0) | 82 (55.0) | 0.79 | 0.54 |

*Supplementary Table 3.* Regression coefficients and p-values for our logistic regression model, with any within-registry outcome discrepancy as the response variable and any within-registry outcome discrepancy as predictor. The predictor did not have missing data.

| **Predictor** | **Predictor Level** | **Any outcome discrepancy** | **No outcome discrepancy** | **Odds Ratio (exponential of regression coefficient)** | **p-Value** |
| --- | --- | --- | --- | --- | --- |
| **Any Within-Registry Discrepancy** | No discrepancy (Intercept) | 95 (42.0) | 131 (58.0) |  |  |
|  | Any discrepancy | 25 (37.9) | 41 (62.1) | 0.84 | 0.55 |

*Supplementary Table 4.* Primary outcome discrepancies reported in the registries at different trial timepoints compared to previous registry entries, and the discrepancies detected in results publications compared to the latest registry entry. This table includes a sensitivity analysis for the registry-publication discrepancies, including only publications with explicitly named primary outcomes (n = 243).

|  | Within-registry discrepancies per trial timepoint (n=1746) | | | |  | Registry-publication discrepancies (n=292; sample) | Registry-publication discrepancies (n=243; sample with only explicitly named outcomes) |
| --- | --- | --- | --- | --- | --- | --- | --- |
|  | Any | recruitment | post-completion | post-publication |  |  |  |
| Discrepancies |  |  |  |  |  |  |  |
| **any** | **393 (22.51%)** | **167 (9.56%)** | **159 (9.11%)** | **131 (7.50%)** |  | **120 (41.10%)**  **[35.40%, 46.98%]** | **86 (35.39%)**  **[29.38%, 41.76%]** |
| major | 142 (8.13%) | 66 (3.78%) | 49 (2.81%) | 36 (2.06%) |  | 54 (18.49%)  [14.21%, 23.43%] | 32 (13.17%)  [9.18%, 18.08%] |
| minor | 318 (18.21%) | 117 (6.70%) | 130 (7.45%) | 110 (6.30%) |  | 75 (25.68%)  [20.77%, 31.10%] | 59 (24.28%)  [19.03%, 30.17%] |
| - changes | 149 (8.53%) | 49 (2.81%) | 61 (3.49%) | 51 (2.92%) |  | 45 (15.41%)  [11.47%, 20.07%] | 34 (13.99%)  [9.89%, 19.00%] |
| - addition/omission | 233 (13.34%) | 78 (4.47%) | 91 (5.21%) | 80 (4.58%) |  | 32 (10.96%)  [7.62%, 15.12%] | 27 (11.11%)  [7.45%, 15.75%] |
| **milestone does not exist** | **5 (0.29%)** | **332 (19.01%)** | **266 (15.23%)** | **945 (54.12%)** |  | **0 (0.00%)**  **[0.00%, 0.00%]** | **0 (0.00%)**  **[0.00%, 0.00%]** |
| **none** | **1348 (77.21%)** | **1247 (71.42%)** | **1321 (75.66%)** | **670 (38.37%)** |  | **172 (58.90%)**  **[53.02%, 64.60%]** | **157 (64.61%)**  **[58.24%, 70.62%]** |
